# Uncovering Distinct Phenotypes of Early Onset Heart Failure in Women

**DOI:** 10.64898/2026.11.04.26367323

**Authors:** Joshua Khorsandi, Yue Yu, Divya Kannan Tinnium, Sandhya Ganesh, Raajameenakshi Alahapphan, Precious Oyinloye, Kamie Mueller, Siamak Ardekani, Casey Overby Taylor, Joseph L. Greenstein, Chiadi E. Ndumele, Erin D. Michos, Noel Bairey Merz, Robert B. Barrett, Anum S. Minhas

**Affiliations:** Department of Biomedical Engineering, Johns Hopkins University, Baltimore, MD; Division of General Internal Medicine, Department of Medicine, Johns Hopkins School of Medicine, Baltimore, MD; Division of Cardiology, Department of Medicine, Johns Hopkins School of Medicine, Baltimore, MD; Barbra Streisand Women’s Heart Center, Cedars-Sinai Smidt Heart Institute, Los Angeles, CA; Division of Biomedical Informatics and Data Science, Department of Medicine, Johns Hopkins School of Medicine, Baltimore, MD

## Abstract

**Background:** Heart failure in younger women is understudied and may reflect heterogeneous pathways and risk not adequately captured by conventional classifications. We aimed to identify clinically distinct phenotypes of early-onset heart failure in women, assess their prognostic significance, and evaluate their transportability in an independent national cohort.

**Methods:** We identified women aged 18-55 years with heart failure using electronic health record data from a large multi-hospital system. A variational deep embedding model incorporating 283 clinical features was used for unsupervised phenotyping. Associations with subsequent inpatient hospitalization, emergency department utilization, and mortality were evaluated using age-stratified Cox models. Phenotype transportability was assessed by both applying the institutionally trained model and by independent de novo clustering as a replication analysis in the All of Us cohort.

**Results:** Among 1,407 young women with heart failure (median age, 45 years), four phenotypes were identified: hypertensive pregnancy (n=494), cardiorenal (n=276), autoimmune (n=394), and cardiometabolic-ischemic (n=243), with a bootstrap adjusted Rand index of 0.65 and silhouette score of 0.45. Compared with the hypertensive pregnancy phenotype, risks of inpatient hospitalization were highest for the cardiorenal (HR 2.86 [95% CI, 2.35–3.49]) and autoimmune (HR 2.25 [95% CI, 1.87–2.70]) phenotypes; mortality risks were similarly elevated (HR 4.65 [95% CI, 3.17–6.82] and HR 4.38 [95% CI, 3.03–6.31], respectively) in these two groups. In All of Us participants (n=682), transferred phenotypes retained their principal clinical characteristics and continued to stratify hospitalization risk. Independent clustering most strongly reproduced the cardiorenal phenotype (Jaccard index, 0.744).

**Conclusions:** Early-onset heart failure in women comprises distinct cardio-obstetric, cardiorenal, autoimmune, and cardiometabolic-ischemic phenotypes with divergent clinical outcomes. These findings support a life-course approach to heart failure classification in women that complements conventional frameworks and incorporates pregnancy history and female-predominant systemic conditions.

**Clinical Perspective:** *What Is New?:*

Unsupervised machine learning identifies distinct clinical phenotypes of heart failure among young women, highlighting heterogeneity in a population that remains understudied.
These phenotypes captured differing cardiometabolic, cardiovascular, and pregnancy-related characteristics, suggesting potentially distinct pathways contributing to heart failure development.
External validation in the All of Us cohort supported external validity and demonstrated the potential for identifying these patterns across diverse healthcare populations.

**What Are the Clinical Implications?:**

Heart failure in young women is clinically diverse, underscoring the importance of comprehensive assessment beyond traditional heart failure classifications, including cardiometabolic risk factors and pregnancy history.
Recognition of distinct clinical phenotypes may help clinicians identify women with differing risk profiles and inform more individualized approaches to cardiovascular evaluation, risk factor management, and longitudinal follow-up.
Further studies are needed to determine whether phenotypes differ in treatment responses and whether phenotype-guided management can improve outcomes.

## INTRODUCTION

Heart failure (HF) remains a major cause of mortality and morbidity among women,^1^ affecting 3 million women in the US alone.^2^ Recent data show that even though overall cardiovascular disease mortality has declined, heart failure mortality rates and hospitalization are rising among women under 55 years.^3,4^ Despite this prevalence, women have traditionally been poorly represented in randomized controlled clinical trials.^5–7^ Moreover, in younger women, pregnancy comprises a common condition, during which women are typically excluded entirely from trials and clinical studies, further reducing representation in evidence generation.

This gap is especially relevant in the context of sex-based differences in epidemiology, pathophysiology, and therapies for heart failure.^8–10^ Biological differences in sex hormones may contribute to modulation of renin-angiotensin-aldosterone, endothelial function, and inflammation.^11^ Women experience unique life-course exposures, such as adverse pregnancy outcomes,^12,13^ premature menopause, and autoimmune diseases, that are not captured by sex-neutral cardiovascular risk stratification tools.^8,14,15^ In addition, although absolute risk of heart failure is higher in older populations, the attributable risk of modifiable risk factors like obesity, hypertension, and diabetes is higher in younger adults (<55 years),^2,16^ prompting the need to study and classify early-onset heart failure. However, this has been frequently limited due to over-reliance on conventional frameworks centered around ejection fraction (EF)^17–20^ and etiology.^21–23^ Unsupervised clustering algorithms have been used to identify novel phenotypes, uncovering latent data-driven groups that may not be apparent with hypothesis-driven models, offering a unique opportunity to better classify heart failure in younger women. However, prior work in phenotype development in heart failure performs clustering on small sample sizes,^24,25^ male-predominant datasets,^24–27^ and cohorts with median age above 65 years,^24–28^ with limited inclusion of younger women. Few prior clustering studies have established external validity of the phenotypes, thereby raising concerns about data driven biases (i.e. clinical practice and data entry patterns) and limiting their potential utility in understanding the disease.

Electronic health records (EHRs) present an opportunity to overcome evidence gaps through a wealth of detailed data, particularly in pregnancy through increased healthcare contact. These data enable real-world evidence generation that may better capture broader population heterogeneity and be replicable at scale for assessing transportability and external validity. Specifically, the Observational Health Data Sciences and Informatics (OHDSI) community has facilitated evidence generation through developing standardized methods for network analysis and federated learning through use of the Observational Medical Outcomes Partnerships (OMOP) Common Data Model (CDM) and standardized vocabularies for data harmonization.^29^ Importantly, CDMs like OMOP provide more usable datasets for reproducing and evaluating external validity of site-specific studies.

In the present study, we aimed to identify and characterize data-driven phenotypes of heart failure among young women, employing unsupervised machine learning for clustering. Secondarily, we aimed to evaluate the prognostic implications of the identified phenotypes for incident outcomes, including hospitalization and urgent visits. To enhance the generalizability and validity of these objectives, we leverage real world EHR data, harmonized using the OMOP CDM, across a large multi-hospital health system, with external validation in the geographically diverse, national All of Us cohort.

## METHODS

### Dataset

Primary analysis was conducted on de-identified longitudinal EHR data, encompassing >2 million patients in the Johns Hopkins Health System (JHHS), from 2016 through 2026. The JHHS OMOP CDM database was used, which organizes patient information into clinical domains, including conditions, drug exposures, measurements, procedures, and observations. The study was approved by the Johns Hopkins Institutional Review Board (IRB00501176). The lead author had access to all data and takes responsibility for integrity of the data and analysis.

### Cohort Inclusion Criteria

The cohort consisted of adult women aged 18-55 years diagnosed with heart failure. The Universal Definition of Heart Failure^30^ was used to determine cohort inclusion, requiring a standardized heart failure diagnosis code (Systematized Nomenclature of Medicine (SNOMED) code = 316139) with either an elevated NT-proBNP level (≥125 pg/mL) or symptoms and signs of heart failure six months prior to the index date, defined as the date of the first heart failure diagnosis code. Patients were excluded if they had no outpatient encounter documented more than six months before the index date.

### Feature Engineering

Patient cohorts were defined using the OHDSI FeatureExtraction R package.^31^ A pre-specified set of 261 features across five domains (conditions, observations, measurements, drugs, procedures) was prepared (**Tables S1-S4**). Descendent concepts were included using the OMOP CDM vocabularies (version 5.4). Patients were randomly assigned to train or test sets at 80% (n=1,125) and 20% (n=282), respectively. Continuous measurements were robustly standardized as (value − median) / IQR and clipped to ±4 to mitigate extreme outliers and measurement errors, with the median and IQR fit exclusively on the training set. Patient conditions, procedures, and observations were included from any time before the index date. Medications were included within two years prior to the index date. Measurement values represented the most recent measurement in the one year window preceding the index date. Using all concept ancestors and a minimum prevalence of 0.01 (calculated from the training set alone), 283 features were ultimately included (**Table S4**).

### Clustering

Given the high-dimensionality of EHR data, we used variational deep embedding (VaDE), an unsupervised clustering approach that integrates dimensionality reduction with probabilistic clustering.^32,33^ VaDE uses a variational autoencoder to learn a lower-dimensional representation of patient characteristics while modeling the latent distribution as a mixture of Gaussian components, allowing patients to be grouped according to multivariable patterns that may not be captured by individual clinical characteristics or linear relationships. We adapted the architecture to accommodate mixed binary and continuous clinical features and incorporated missingness masks to distinguish unobserved measurements from observed values.

Patients were assigned to clusters according to the posterior probability of membership in the learned Gaussian mixture components. Clinical labels were subsequently assigned through expert clinical interpretation of cluster-specific feature enrichment and descriptive characteristics. Further details on the clustering architecture are presented in the supplementary methods.

### Hyperparameters and Evaluation

Patients were randomly partitioned into a training (80%, n=1,125) and hold-out test set (20%, n=282). Model weights were fit on the training set only, and the scaling and imputation parameters for continuous measurements were likewise estimated from training data alone. Model hyperparameters, including the dimensionality of the latent representation and number of Gaussian mixture components, were optimized using

Bayesian optimization in the Optuna framework.^33^ Hyperparameter selection considered two competing objectives: minimizing the variational autoencoder loss on the hold-out set and maximizing the average silhouette score. Candidate configurations were evaluated along the Pareto frontier to identify solutions balancing representation quality and cluster separation. The held-out set was also used to determine early stopping during training.

Reconstruction performance of the final model was evaluated on the hold-out set separately for binary and continuous features. Because binary features were sparse (7.0% of entries positive), accuracy was not informative; we instead report the area under the receiver operating characteristic curve (AUROC), the area under the precision–recall curve (AUPRC), and precision, recall, and F1 score for positive values, with reconstructed probabilities of 0.5 or greater classified as positive. Continuous features were evaluated on observed values only, using mean absolute error (MAE), root mean squared error (RMSE), the coefficient of determination (R²), and Pearson correlation, and were compared with a baseline that predicts each feature’s training-set mean. Metrics were computed both pooled across all entries and as the median across features. To assess the robustness and validity of cluster assignments, we performed 100 bootstrap-based clustering evaluations and quantified agreement using the adjusted Rand index (ARI). Final model hyperparameters and metrics are reported in **Table S5**.

### Posthoc Clustering Explanations

We trained a supervised eXtreme Gradient Boosting (XGBoost) model^34^ to predict cluster labels from the unsupervised model using the original 283-feature dataset. SHapley Additive exPlanations (SHAP) analysis was then used to quantify feature importances and contribution of individual features as previously described.^35^ The XGBoost model was trained on a 1,126 patient randomized dataset with evaluation metrics derived from the remaining 281-patient test set. The final F1 score is reported as the size weighted average across all clusters. Additionally, in order to descriptively characterize cluster-specific feature enrichment, we calculated log odds ratios for binary features and standardized mean differences (SMDs) for continuous features.

### Risk Assessment and Survival Analysis

We evaluated the association between cluster membership and subsequent healthcare utilization using Kaplan-Meier survival estimates and Cox proportional hazards regression. Four outcomes were examined: time from the heart failure index date to first inpatient admission, first emergency department (ED) visit, a composite first occurrence of either event, and mortality. Follow-up extended from the heart failure index date to either the first qualifying event or last observed clinical encounter.

Cox proportional hazards models were stratified by age quartiles determined at the heart failure index date, with the largest cluster serving as the reference group.

Proportional hazards assumptions were assessed using Schoenfeld residuals. Kaplan-Meier curves were generated to estimate event-free survival over the follow-up period.

In a secondary analysis, patients were classified by heart failure subtype using ICD-10-CM diagnoses mapped to OMOP concepts (**Table S6**). Systolic or combined heart failure codes defined heart failure with reduced ejection fraction (HFrEF), while diastolic heart failure or right-sided heart failure defined heart failure with preserved ejection fraction (HFpEF). Patients without qualifying subtype codes were excluded for subtype analyses. Age-adjusted Cox models evaluated cluster-associated mortality and subsequent hospitalization within subtypes, with pooled estimates stratified by subtype.

### External Validation and Independent Replication in All of Us

We evaluated the external validity and reproducibility of JHHS-derived heart failure phenotypes using the independent All of Us cohort through two complementary approaches: external model transfer and independent clustering. For both approaches, we applied the same cohort definition and feature extraction across JHHS and All of Us cohorts. All of Us data are standardized to the OMOP CDM (v5.4). Cohort extraction was performed via BigQuery queries against the All of Us Controlled Tier Curated Data Repository. Features represented in fewer than 10 patients were dropped. Missingness was handled by imputing the JHHS training mean when a measurement had no recorded value.

For independent replication, a separate VaDE model was trained. The number of clusters was fixed to match JHHS, while remaining hyperparameters were optimized following the primary (JHHS) analysis approach (**Table S7**). Patients were assigned to the phenotype with the highest posterior membership probability. Cluster stability was evaluated across five random seeds using an 80/20 training-testing split. Agreement between independently derived and transferred cluster assignments was assessed using ARI and cluster-level Jaccard similarity. Survival analysis was similarly performed in All of Us for subsequent hospitalization. Mortality analyses were not repeated in All of Us, given prevalence limiting stable model fit.

## RESULTS

The final cohort was comprised of 1407 young women (median age 45 [IQR 37-52] years) with heart failure (**Figure 1**). The majority of patients identified as African American (n=781, 55.5%) followed by White (n=490, 34.8%). Obesity was common, with median BMI 31.7 kg/m² and 52.0% with an obesity diagnosis. Blood pressure was generally within the normal range (median 114/65 mmHg). Median hemoglobin was low at 10.3 g/dL, and median eGFR was 82.8 mL/min per 1.73 m². Anemia was the most frequent comorbidity (67.0%), followed by hyperlipidemia (37.1%), type 2 diabetes (33.8%) and chronic kidney disease (27.9%). Pregnancy-related conditions were also prevalent, with 9.5% with a high-risk pregnancy and 4.3% with pre-eclampsia (Table 1, **Table S8**). Following VAE fitting, the optimized model used 10 latent dimensions and partitioned patients into four clusters. The resulting clustering solution maintained a bootstrap ARI of 0.65±0.02 and an average silhouette score of 0.45 across all clusters. On the hold-out test set, the model reconstructed binary features with a cluster size weighted average AUROC of 0.80 and an AUPRC of 0.24 (against a positive prevalence of 0.07). For continuous features, it achieved an MAE of 0.63 and R² of 0.23, compared with an MAE of 0.71 for the training-mean baseline (**Table S5**).

**Figure 1.**
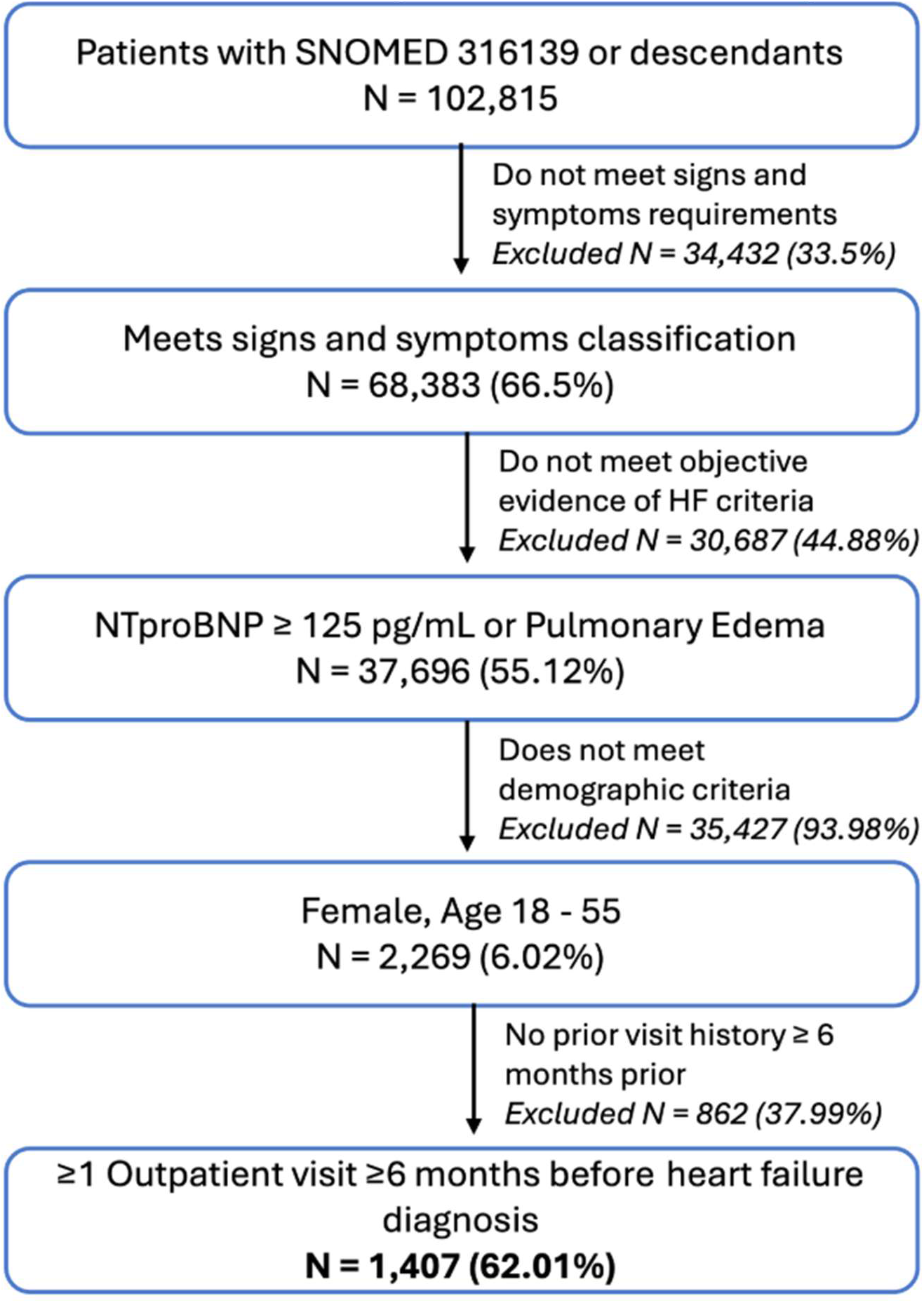
Cohort Definition and Study Inclusion for Johns Hopkins Health System. **Figure 1 Legend**:The Johns Hopkins Health System cohort includes women aged 18-55 years who meet the study criteria: signs and symptoms of heart failure plus either pulmonary edema or elevated natriuretic peptide. To establish prior history, patients also needed at least one outpatient visit at least six months prior to the heart failure diagnosis date.

**Table 1.**
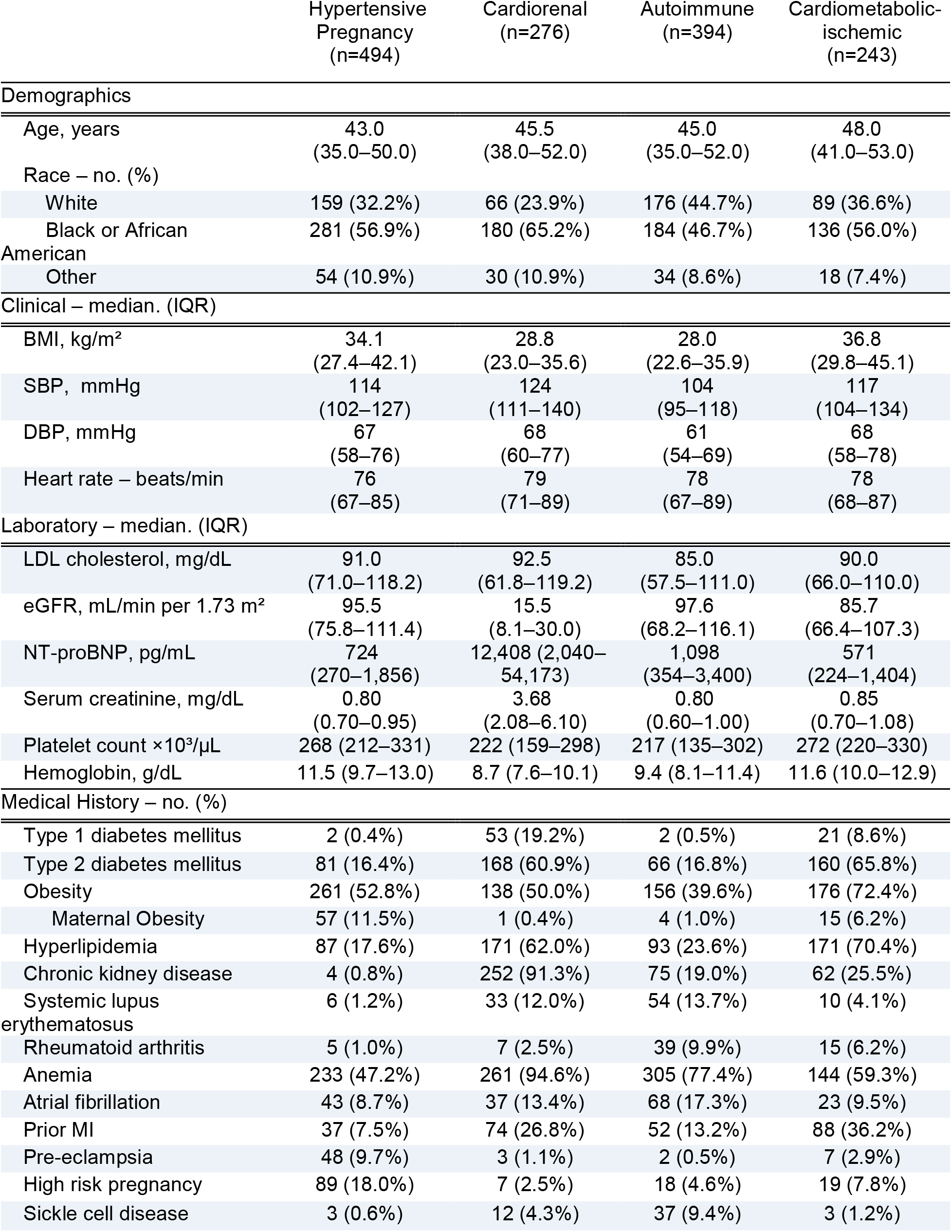
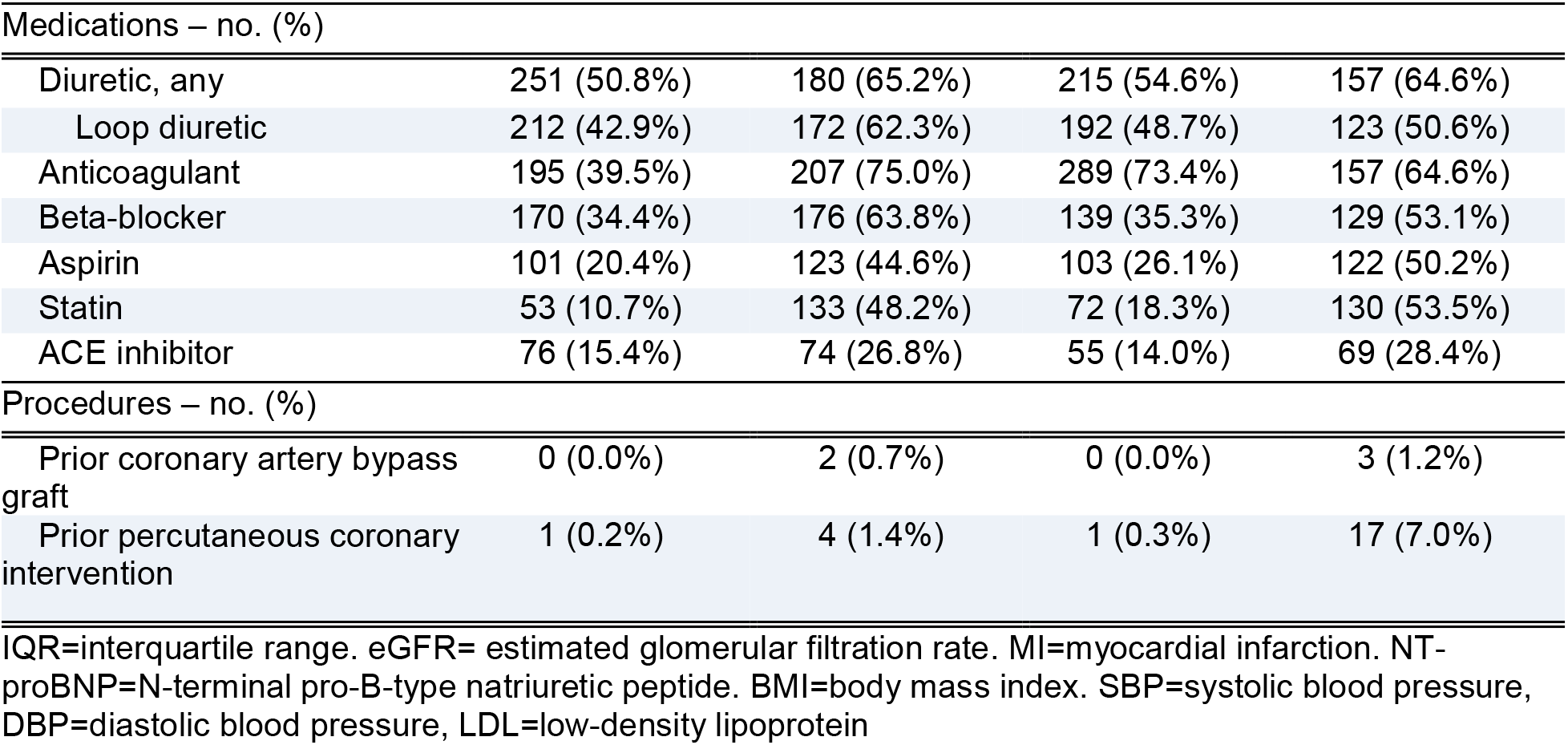
Clinical characteristics across phenotypes of heart failure in the JHHS cohort.

### Cluster Characterization

Following expert review of cluster characteristics by a cardiologist, clusters were determined as: hypertensive pregnancy (n=494), cardiorenal (n=276), autoimmune (n=394), and cardiometabolic-ischemic (n=243).

The hypertensive pregnancy cluster was the largest and youngest (median age 43.0 years, IQR [35, 50]), with the second highest prevalence of obesity (n=261, 53% of the cluster). This cluster was enriched for pregnancy-related outcomes, including preeclampsia (n=48, 10% of cluster) and high-risk pregnancy (n=89, 18% of cluster).

The cardiorenal cluster was primarily characterized by reduced renal function, with lower eGFR (median 15.5 mL/min per 1.73 m ^2^ [IQR 8.1-30.0]) and elevated serum creatinine (median 3.68 mg/dL, IQR [2.08, 6.10]). The majority of the cluster (91%) had a diagnosis of chronic kidney disease.

The autoimmune cluster had the highest proportion of patients with autoimmune disease, particularly systemic lupus erythematosus (n=54, 14% of cluster). This cluster also had the greatest proportion of individuals with pulmonary hypertension and sickle cell disease.

The cardiometabolic-ischemic cluster had the highest body mass index (BMI) (median 36.8 kg/m^2^, IQR [29.8, 45.1]) and within cluster prevalence of type 2 diabetes (66%), obesity (72%), and myocardial infarction (36%).

Distributions of key features and relative prevalence within each cluster are displayed in **Figure 2, panels A-B**.

**Figure 2.**
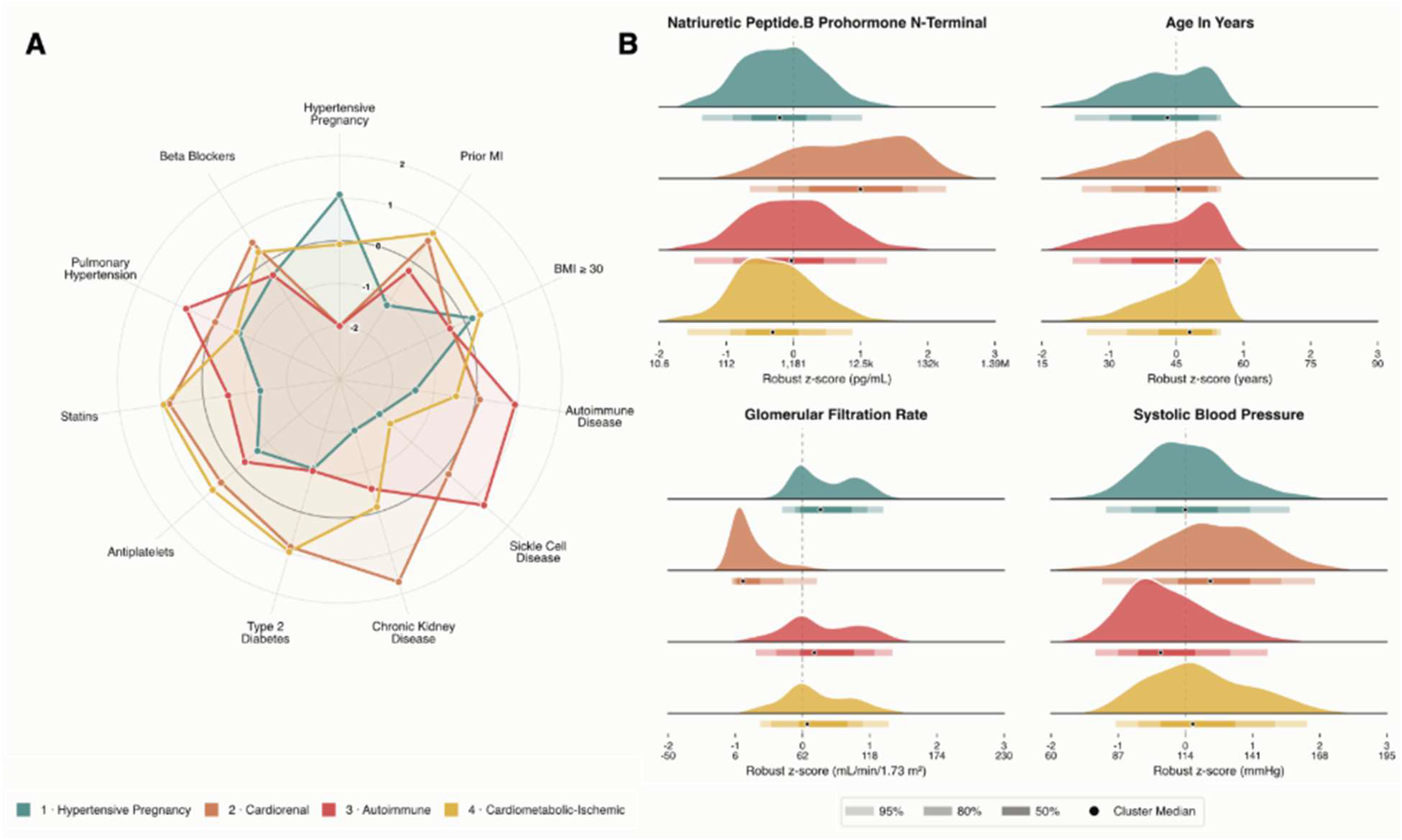
Clinical and Laboratory Characteristics across Heart Failure Phenotypes. **Figure 2 Legend.** Panel A demonstrates cluster identities defined by condition and drug enrichment. Concentric rings represent log2 odds ratio per condition or drug, per cluster. Inner bolded ring represents no enrichment over cohort (log2 odds ratio of 0). Panel B displays per cluster measurement distributions. Ridgelines represent per-cluster patient density vs. robust z-score for notable continuous features.

### Post-hoc Model Explanation and Validation

Feature contribution analysis, using XGBoost to predict individual cluster membership, achieved an F1 score of 0.77 and an average, one-against-all, average AUC-ROC of 0.94. Cluster AUC-ROC ranged from 0.90 for the cardiometabolic-ischemic cluster to 0.99 for the cardiorenal cluster. Using the mean absolute SHAP values across all patients, we noted that the most influential feature was serum creatinine, which predicted mostly for cardiorenal cluster membership (**Figure S1**). Feature importance was consistent with cluster identities as descriptively determined.

Per-patient per-feature SHAP values were also computed per cluster for the patient with the highest assignment probability (**Figure 3**). The features most influencing cluster assignment were prior preeclampsia diagnosis and prior ethinyl estradiol prescription for hypertensive pregnancy cluster, high creatinine and end-stage renal disease diagnosis for the cardiorenal cluster, pancytopenia with depressed platelet counts for the autoimmune cluster, and history of aortocoronary bypass graft and ST segment elevation myocardial infarction for the cardiometabolic-ischemic cluster.

**Figure 3.**
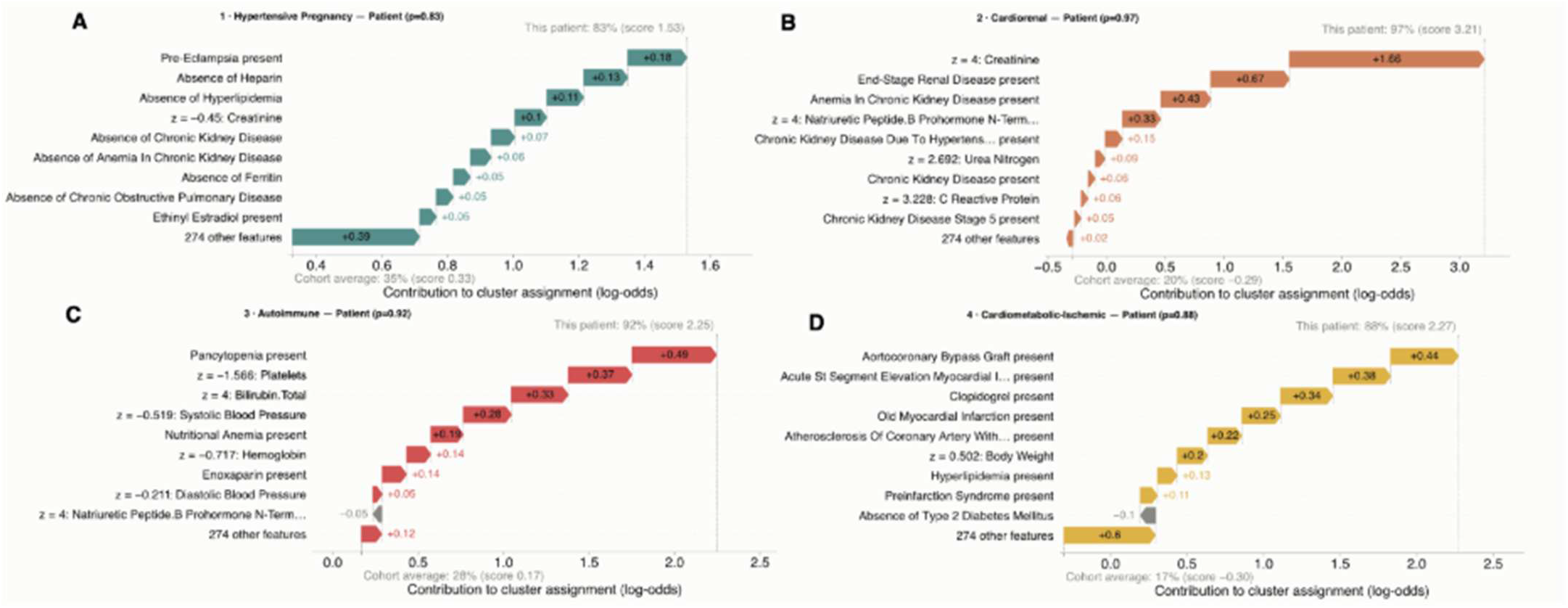
Additive value of key features on model cluster assignment. **Figure 3 Legend.** Per feature SHAP values for representative patients across clusters (panels A – D), with model confidence p=0.83 (A, Hypertensive Pregnancy), 0.97 (B, Cardiorenal), 0.92 (C, Autoimmune), and 0.88 (D, Cardiometabolic-Ischemic).

### Stratified Risk for Rehospitalization

Hazard analyses were conducted using the hypertensive pregnancy cluster as reference. 807 inpatient all-cause subsequent hospitalization events occurred among 1406 patients (57.4%), over median 1.2 years of follow-up. The cardiorenal cluster had the greatest HR at 2.86 (95% CI 2.35-3.49, p<0.001), followed by autoimmune at 2.25 (95%CI 1.87-2.70, p<0.001) and cardiometabolic-ischemic at 1.55 (95% CI 1.25-1.92, p<0.001). For ED only, 590 events occurred among 1406 patients (42.0%). The cardiometabolic-ischemic cluster had the greatest risk with HR 1.50 (95%CI 1.20-1.89, p<0.001), followed by cardiorenal at 1.33 (95%CI 1.06-1.66, p= 0.013), and autoimmune at 0.80 (95%CI 0.64-1.00, p=0.052) (**Figure 4A**). For the composite inpatient and ED outcome, 959 events occurred among 1406 patients (68.2%). The cardiorenal cluster had HR 2.00 (95% CI 1.67-2.39, p<0.001), followed by autoimmune at 1.52 (95% CI 1.29-1.79, p<0.001) and cardiometabolic-ischemic at 1.38 (95% CI 1.13-1.67, p<0.001) (**Figure 4A, Figure S2**).

**Figure 4.**
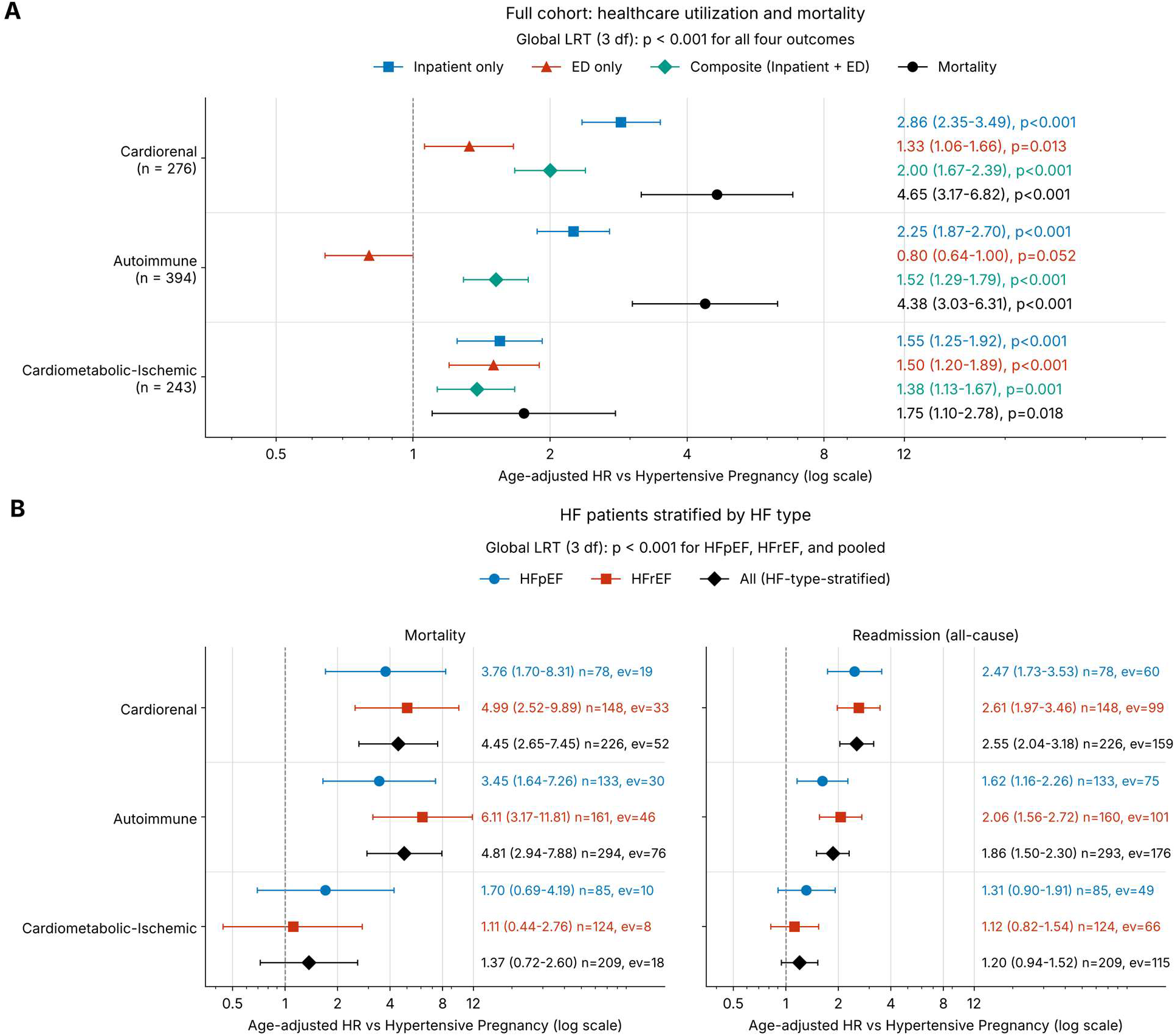
Healthcare utilization and mortality by phenotype cluster, overall and stratified by heart failure type. **Figure 4 Legend.** Panel A shows age-adjusted Cox hazard ratios for healthcare utilization and mortality by cluster, relative to the Hypertensive Pregnancy cluster. Hazard ratios are shown with 95% confidence intervals on a log scale; the dashed line marks HR = 1. Panel B shows the subset of patients with a heart failure, stratified by diagnosis-based classifications for HFpEF or HFrEF, with mortality (left) and all-cause readmission (right) estimated separately for HFpEF (circles), HFrEF (squares), and the HF-type-stratified pooled model (diamonds). Global likelihood ratio tests (3 df) are shown above each panel. Hazard ratios are plotted on a log scale; n = patients, ev = events.

### Mortality Analysis

A total of 279 deaths occurred (19.8% of the cohort). Compared to the reference hypertensive pregnancy cluster, the cardiorenal cluster had HR 4.65 (95% CI 3.17-6.82, p<0.001), the autoimmune cluster had HR 4.38 (95% CI 3.03-6.31, p<0.001) and the cardiometabolic-ischemic cluster had HR 1.75 (95% CI 1.10-2.78, p=0.018). Kaplan-Meier mortality curves are shown in **Figure S3**. When stratified by heart failure type, the elevated risks of mortality and all-cause subsequent hospitalization in the cardiorenal and autoimmune phenotypes were consistent among patients with HFpEF and HFrEF (**Figure 4B**).

### Validation in the All of Us Cohort

The All of Us cohort included 682 young women with heart failure, with 266 of 283 JHHS-derived features (94%) available for analysis (**Figure S4**). Application of the JHHS-trained model to All of Us yielded four clusters that broadly preserved the clinical characteristics observed in the derivation cohort, although their relative prevalence and degree of feature enrichment differed (**Table S9**). The transferred solution had a mean silhouette score of 0.31.

The hypertensive pregnancy phenotype retained enrichment for pregnancy-induced hypertension and preeclampsia; the cardiorenal phenotype remained characterized by chronic kidney disease, end-stage renal disease, anemia of chronic kidney disease, and reduced eGFR; the autoimmune phenotype retained enrichment for systemic lupus erythematosus, pulmonary hypertension, sickle cell disease, and pulmonary embolism; and the cardiometabolic-ischemic phenotype remained enriched for obesity, diabetes, and hyperlipidemia (**Figure 5; Figure S5**).

**Figure 5.**
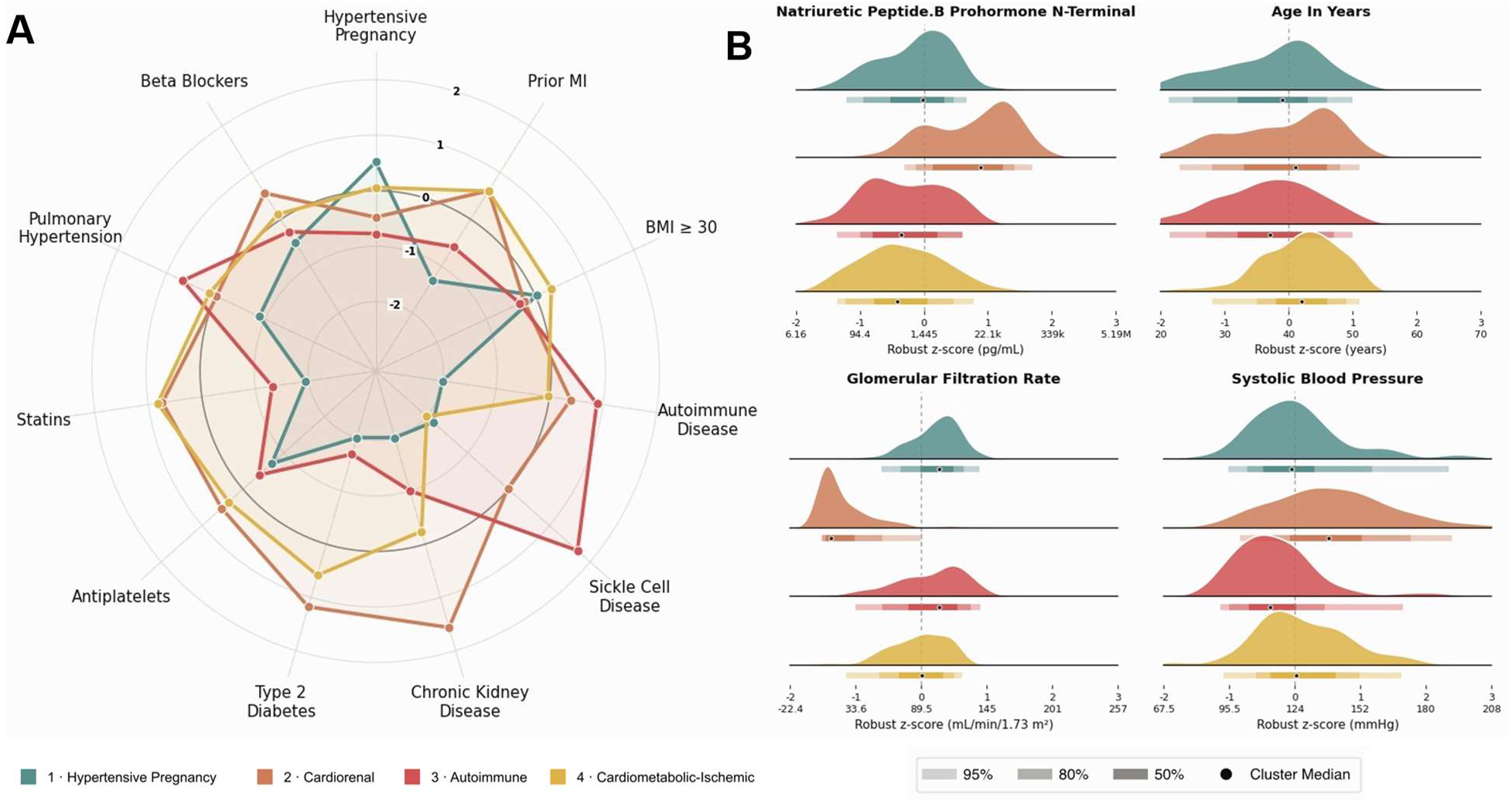
Phenotypic characterization of JHHS-transferred clusters in All of Us. **Figure 5 Legend.** Figure shows the phenotypic characterization of JHHS transferred clusters in All of Us. Hypertensive Pregnancy Cluster (n = 213); Cardiorenal Cluster (n = 150); Autoimmune Cluster (n = 138); and Cardiometabolic-Ischemic (n = 181). Colors identify the same clusters across all panels. Panel A shows cluster identities as characterized by condition and drug enrichment, expressed as log₂ prevalence ratios relative to the overall cohort. Panel B displays per-cluster distributions of NT-proBNP, age, glomerular filtration rate, and systolic blood pressure, displayed on robust z-score scales. Black dots indicate cluster medians, and shaded bars represent central 50%, 80%, and 95% intervals.

In the independent de novo clustering analysis, correspondence was strongest for the cardiorenal phenotype, with 85% overlap with the transferred cardiorenal cluster (Jaccard index, 0.744), whereas pregnancy-related, autoimmune, and cardiometabolic features were distributed less consistently across the remaining clusters (ARI 0.234; adjusted mutual information 0.286; **Table S10**; **Figures S6-S7**).

Despite these differences in de novo cluster structure, the transferred phenotypes continued to inform subsequent hospitalization in All of Us. Compared with the cardio-obstetric phenotype, the cardiorenal phenotype had the highest risk of subsequent inpatient hospitalization (HR 1.71 [95%CI 1.24-2.36]), while the cardiometabolic-ischemic phenotype was associated with increased risk of inpatient hospitalization (HR 1.62 [95%CI 1.17-2.23]), ER utilization (HR 1.60 [95%CI 1.22-2.11]), and the composite outcome (HR 1.58 [95%CI 1.22-2.06]) (**Figures S8-S9**). Detailed characteristics of the transferred phenotypes are provided in Table S9 and Figure 5.

## DISCUSSION

In this real-world study of young women with heart failure, unsupervised learning of the EHR-identified four clinically distinct phenotypes characterized by hypertensive pregnancy, cardiorenal, autoimmune, and cardiometabolic-ischemic features. Three findings are particularly notable. First, two of the four phenotypes were organized around conditions that uniquely or disproportionately affect women, specifically hypertensive disorders of pregnancy and autoimmune disease. These conditions have rarely emerged as defining characteristics in prior heart failure phenotyping studies conducted largely in older or mixed-sex populations.^36–39^ Second, phenotype membership was associated with divergent risks of subsequent hospitalization and mortality. Third, distinct phenotypes were identified with differing risks for subsequent hospitalization and mortality, despite similar distribution of HFrEF and HFpEF, suggesting that EF alone may incompletely characterize early-onset heart failure in women. Finally, distinguishing features among clusters remained present with transfer of the institutionally derived model to the multicenter All of Us cohort. Together, these findings suggest that heart failure in younger women may represent a clinical syndrome arising through several life-course and systemic disease pathways, rather than a single disease process.

The emergence of a distinct hypertensive pregnancy phenotype is particularly important. This was the largest and youngest group and was enriched for high-risk pregnancy and obesity. Hypertensive disorders of pregnancy are increasingly recognized as markers of premature cardiovascular disease, and recent longitudinal studies have demonstrated associations between adverse pregnancy outcomes and subsequent heart failure.^12^ However, pregnancy history has rarely been incorporated into prior heart failure phenotyping studies.^36–38^ Women in this group experienced the lowest subsequent hospitalization and mortality risk compared with the other phenotypes. However, this should not be interpreted as indicating low absolute cardiovascular risk, as all participants had established heart failure. This phenotype may identify women in whom heart failure occurs earlier in the life course with unique, modifiable risk factors for developing renal, inflammatory, or ischemic comorbidities. Longitudinal studies beginning before heart failure onset will be needed to determine whether this group represents an earlier stage along a cardiometabolic-HFpEF trajectory or a mechanistically separate pathway to heart failure.

The autoimmune phenotype represents another potentially important pathway to early heart failure in women. This cluster was enriched for systemic lupus erythematosus, rheumatoid arthritis, pulmonary hypertension, sickle cell disease, anemia, and hematologic abnormalities. Women bear a disproportionate burden of many systemic autoimmune diseases, and growing evidence links chronic immune activation, endothelial dysfunction, and myocardial inflammation to HFpEF, independent of traditional cardiovascular risk factors.^40^ Although we cannot establish causal mechanisms in the current study, these findings support inclusion of autoimmune history into heart failure risk assessment and phenotyping in women.

The cardiorenal cluster and cardiometabolic-ischemic clusters are consistent with prior heart failure phenotyping studies.^36,38^ The cardiorenal phenotype was characterized by renal dysfunction, whereas the cardiometabolic-ischemic phenotype had the highest prevalence of obesity, type 2 diabetes and coronary artery disease. Notably, prior clustering studies in older heart failure populations have often identified broader multimorbidity or metabolic-vascular groups in which renal, metabolic, microvascular dysfunction, and vascular disease coexist.^36^ The separation of these conditions into renal-dominant and ischemic-metabolic phenotypes in the present cohort may reflect the younger population studied, before extensive age-related multimorbidity obscures distinct disease pathways, as well as the broader clinical feature space used for clustering.

External analyses replicating the cluster phenotypes in All of Us provide additional insight. When the institutionally-trained model was applied directly, the principal clinical features of all four phenotypes remained consistent, and clusters remained distinct with observable differences in hospitalization risk. The cardiorenal phenotype presented with higher inpatient admission risk and the cardiometabolic-ischemic phenotype demonstrated increased inpatient, ER, and composite utilization. Together, these analyses support the transportability and prognostic relevance of the JHHS-derived phenotypes, although their partial correspondence with independently derived clusters suggests that the underlying cluster structure varies across populations. This is important because many prior heart failure clustering studies have been derived and evaluated within a single cohort, limiting inference regarding generalizability.

This study did not completely reproduce the JHHS derived clusters in All of Us through de novo analysis. Correspondence was strongest for the cardiorenal phenotype, whereas pregnancy-related, autoimmune, and metabolic features were distributed less consistently across independently derived clusters. This distinction between transportability and exact reproducibility is informative. Severe renal dysfunction may constitute a more stable axis of heart failure heterogeneity across populations, while hypertensive pregnancy and autoimmune phenotypes may be more sensitive to differences in population composition, health-system contact, coding practices, reproductive history capture, or feature prevalence.

Several limitations should be considered. First, the study was observational and based on routinely collected EHR data. Therefore, diagnoses and exposures were dependent on clinical documentation and health-system contact, and measurements may reflect healthcare utilization patterns instead of true prevalence. Second, the clusters should be interpreted as data-derived phenotypes rather than causal disease subtypes. Third, unmeasured social, environmental, and structural determinants of health may contribute importantly to early-onset heart failure and should be incorporated into future phenotyping studies. Finally, more complete echocardiographic characterization was not available but may be important for biological refinement of these phenotypes.

Overall, these findings have implications for conceptualization of heart failure in young women. Current classification systems appropriately emphasize EF given benefit for evidence-based heart failure therapies; however, the present results suggest that a second dimension of classification, i.e. one incorporating how patients arrive at heart failure across the life course, may be useful. Pregnancy-related vascular disease, autoimmune disease, severe renal dysfunction, and premature cardiometabolic or ischemic disease may reflect different pathways converging on early heart failure presentation. Such a framework may be particularly relevant for prevention. Future studies should investigate potential biological mechanisms underlying the differences among clusters and whether groups may benefit from individualized treatment strategies.

## Data Availability

Primary analysis was performed in the Johns Hopkins Health System electronic health record. Patient privacy rules prevent us from sharing data directly but code can be made available upon request. All of Us data can be directly accessed by approved investigators from the study site with appropriate permissions.

## Acknowledgements

We would like to thank the patients of the Johns Hopkins Health System and participants of the All of Us cohort for access to their de-identified data.

## Non-standard Abbreviations and Acronyms

HF: Heart Failure
EF: Ejection Fraction
EHR: Electronic Health Record
OMOP: Observational Medical Outcomes Partnership
OHDSI: Observational Health Data Sciences and Informatics
CDM: Common Data Model
SNOMED: Systematized Nomenclature of Medicine
VaDe: Variational Deep Embedding
XGBoost: Extreme Gradient Boosting
SHAP: SHapley Additive exPlanations
SMD: Standardized Mean Difference
ARI: Adjusted Rand Index
ER: Emergency Room
JHHS: Johns Hopkins Health System
HFrEF: Heart Failure with Reduced Ejection Fraction
HFpEF: Heart Failure with Preserved Ejection Fraction
BMI: Body Mass Index
HR: Hazard Ratio

